# Fuzzy Logic Model for Healthcare Resource Allocation Optimization in Diabetes Care: Analysis of Brazilian Unified Health System (SUS) Administrative Databases

**DOI:** 10.64898/2025.11.30.25341309

**Authors:** Luís Jesuino de Oliveira Andrade, Luisa Correia Matos de Oliveira, Luanna Lopes da Silva Ramos, Gabriela Correia Matos de Oliveira, Larissa Morgana Carvalho Santos, Luís Matos de Oliveira

## Abstract

**Introduction:** Healthcare systems worldwide face mounting challenges in resource allocation amid the rising burden of chronic diseases and persistent budgetary constraints. In Brazil, the Unified Health System (SUS) must deliver comprehensive care for people with diabetes mellitus (DM) while optimizing scarce resources. Fuzzy inference approaches provide a flexible framework capable of accommodating these complexities; however, evidence regarding their application to national-level diabetes care planning remains scarce.

**Objective:** To develop and validate a fuzzy logic-based optimization model to identify more effective, equitable, and outcome-oriented resource allocation strategies for diabetes care within SUS.

**Methods:** We conducted a retrospective cross-sectional study utilizing DATASUS, SIH-SUS, and Hiperdia registries spanning January 2015 to December 2024 across 5,570 Brazilian municipalities, and constructed a hierarchical Mamdani-type fuzzy inference system incorporating epidemiologic, economic, clinical, and structural indicators. The model was calibrated using historical data, validated through technical, empirical, and expert assessment, and embedded within a multi-objective optimization framework to evaluate alternative investment scenarios across varied budget constraints.

**Results:** The integrated dataset comprised 8,347,219 diabetes-related hospitalizations. The fuzzy inference system demonstrated 97.3% coverage and outperformed conventional approaches with mean absolute percentage error of 12.4% for expenditure predictions. Under baseline conditions, the model recommended increasing primary care investments from 31.2% to 42.7% while reducing tertiary hospital care from 38.4% to 28.9%. These reallocations predicted 8.4% improvement in glycemic control, 12.7% reduction in hospitalizations, and 6.2% mortality decrease over five years. Geographic analysis identified 847 highest-priority municipalities requiring targeted intervention.

**Conclusion:** Fuzzy logic-based optimization demonstrates substantial potential for enhancing diabetes care efficiency through strategic reallocation prioritizing primary care expansion and equity-focused interventions in underserved regions.

## INTRODUCTION

Fuzzy logic systems have emerged as powerful computational frameworks for addressing uncertainty and imprecision inherent in complex healthcare decision-making processes.^1^ Unlike traditional binary logic, fuzzy set theory accommodates the gradual transitions and ambiguous boundaries characteristic of real-world clinical scenarios, enabling more nuanced modeling of resource allocation challenges.^2^ The mathematical foundations of fuzzy inference systems, originally developed by Zadeh in 1965, provide precisely this capability through linguistic variables and membership functions that mirror human expert reasoning patterns.^3^ Recent applications in healthcare resource management have demonstrated that fuzzy logic approaches can outperform conventional optimization techniques, particularly when dealing with incomplete data sets or conflicting stakeholder priorities.^4^

Brazil’s Unified Health System (SUS) faces mounting pressure to deliver comprehensive diabetes care to approximately 16.8 million affected individuals within constrained budgetary frameworks.^5^ The Ministry of Health allocates substantial resources through Department of Informatics of the Unified Health System (DATASUS) administrative databases, yet inefficiencies persist in translating expenditure into improved patient outcomes.^6^ Diabetes-related hospitalizations recorded in the Hospital Information System (SIH-SUS) system have increased 23% over the past decade, with associated costs surpassing R$3.9 billion annually, highlighting urgent needs for evidence-based resource allocation strategies.^7^ Traditional optimization models applied to SUS data often fail to capture the heterogeneous nature of diabetes care requirements across Brazil’s diverse geographic and socioeconomic landscape.^8^ The Ministry of Health program dedicated to the registration and continuous monitoring of patients with hypertension and diabetes mellitus (Hiperdia) registry, containing longitudinal data on over 7 million diabetic patients, remains underutilized for predictive analytics that could guide strategic investment decisions in prevention, treatment, and complication management programs.^9^

Despite growing recognition that intelligent resource allocation could substantially improve diabetes care efficiency within SUS, current approaches lack sophisticated decision-support mechanisms that accommodate the multidimensional uncertainty characterizing healthcare systems. Existing literature demonstrates successful fuzzy logic applications in clinical diabetes management but provides limited evidence regarding its utility for macro-level health system optimization using administrative databases.^10^ No published studies have systematically applied fuzzy inference systems to integrate DATASUS cost data with clinical outcomes from SIH-SUS and Hiperdia registries.

This study aims to develop and validate a comprehensive fuzzy logic model for optimizing healthcare resource allocation in diabetes care within Brazil’s Unified Health System, utilizing integrated analysis of DATASUS, SIH-SUS, and Hiperdia databases to identify efficient investment strategies that maximize population health outcomes while respecting budgetary constraints.

## METHODS

### Study Design and Data Sources

This retrospective cross-sectional study employed secondary data analysis from three interconnected Brazilian SUS administrative databases spanning January 2015 to December 2024. Data were extracted from DATASUS, SIH-SUS, and Hiperdia registries. The study protocol received exemption from ethics committee review as it utilized exclusively de-identified, publicly available aggregate data in accordance with Brazilian National Health Council Resolution 510/2016. All procedures adhered to RECORD guidelines for transparent reporting of research using routinely collected health data.

### Database Integration and Preprocessing

We integrated data sources using deterministic linkage algorithms based on municipal health region identifiers (IBGE codes) and hospital registration numbers (CNES codes). From DATASUS, we extracted annual budgetary allocations for diabetes-related services across all 5,570 Brazilian municipalities. SIH-SUS provided hospitalization records for diabetes-related admissions (ICD-10 codes E10-E14), including costs, complications, and mortality outcomes. Hiperdia contributed patient-level information on diabetes prevalence, HbA1c control rates, and comorbidity profiles aggregated at municipal levels.

Data quality assessment involved systematic evaluation of completeness and consistency. Records with missing critical variables (>30% incomplete) or implausible values were excluded from specific analyses. Geographic coordinates and sociodemographic indicators from IBGE supplemented the health databases to enable spatial analysis and adjustment for regional heterogeneity.

### Fuzzy Logic Model Development

#### Conceptual Framework and Input Variables

We developed a hierarchical Mamdani-type fuzzy inference system incorporating five primary domains: (1) epidemiological burden; (2) healthcare resource utilization; (3) clinical outcomes; (4) economic indicators; and (5) structural capacity. Input variables were selected through modified Delphi consensus involving Brazilian endocrinologists, health economists, and public health administrators. Three iterative rounds achieved consensus (≥80% agreement) on 24 key variables distributed across domains.

#### Fuzzification and Membership Functions

For each variable, we defined three to five fuzzy sets representing linguistic terms (e.g., “very low,” “low,” “moderate,” “high,” “very high”) with corresponding membership functions. Shapes (triangular, trapezoidal, or Gaussian) were determined through empirical data distribution analysis. Fuzzification parameters were calibrated using 2015-2019 data, with 2020-2024 reserved for validation. For instance, diabetes prevalence was fuzzified using five overlapping trapezoidal functions: very low (3-6%), low (5-8%), moderate (7-10%), high (9-13%), and very high (12-16%). Cost variables underwent logarithmic transformation before fuzzification. All membership functions were normalized to the [0,1] interval.

#### Fuzzy Rule Base Construction

We constructed 156 if-then fuzzy rules structured in three hierarchical levels addressing domain-specific relationships, cross-domain interactions, and budget constraint scenarios. Rule weights were assigned through analytic hierarchy process methodology with expert pairwise comparisons. Consistency ratios remained below 0.10, indicating acceptable logical consistency. Conflict resolution among contradictory rules utilized weighted averaging methods.

#### Inference Engine and Defuzzification

The fuzzy inference process employed Mamdani’s minimum implication with maximum aggregation. We implemented the system using Octave 8.4.0 with the Fuzzy Logic Toolkit. The system processed municipal-level data iteratively, generating fuzzy outputs for six strategic investment categories: primary care expansion, specialist services, medication programs, complication prevention, health technology, and patient education. Defuzzification employed the centroid method to convert fuzzy outputs into crisp numerical values representing optimal resource allocation percentages.

#### Optimization Framework

We formulated resource allocation as multi-objective constrained optimization with three objectives: (1) maximize population health outcomes; (2) minimize healthcare expenditure per capita; and (3) maximize equity in care access. Constraints included budget limitations, minimum investment thresholds, and capacity restrictions. The framework integrated fuzzy recommendations with genetic algorithms implemented in Python 3.11 using DEAP library (Distributed Evolutionary Algorithms in Python). The Genetic Algorithm employed tournament selection, uniform crossover (probability=0.8), and polynomial mutation (probability=0.2) across 200-solution populations evolved over 500 generations. Non-dominated Sorting Genetic Algorithm II implementation identified Pareto-optimal solutions.

#### Validation and Performance Assessment

Model validation employed technical, empirical, and expert validation approaches. Technical validation assessed completeness, consistency, and continuity, demonstrating 97.3% coverage of observed input combinations. Empirical validation utilized 2020-2024 out-of-sample testing, comparing model recommendations with actual investments and outcomes. Performance metrics included mean absolute percentage error, AUROC for high-need municipality identification, and Spearman correlations between predicted and observed improvements. We benchmarked against linear programming, machine learning approaches (random forest, gradient boosting), and historical allocation patterns.

Expert validation convened two independent healthcare policy specialists reviewing 50 randomly selected recommendations using structured evaluation forms. Inter-rater reliability was assessed through Fleiss’ kappa statistics.

#### Scenario and Sensitivity Analyses

Comprehensive scenario analyses explored model behavior under budget variations (−20%, baseline, +20%), changing epidemiological trajectories, and policy interventions. Sensitivity analyses systematically varied input parameters and fuzzy components. One-at-a-time analysis altered each variable across feasible ranges. Sobol indices decomposed output variance into contributions from individual inputs and interactions. Structural sensitivity modified membership functions, rule weights, and aggregation operators.

#### Statistical Analysis

Analyses were performed using R 4.3.2 and Python 3.11 with scikit-fuzzy, scikit-learn, and NetworkX libraries. Continuous variables were reported as means ± SD or medians with IQR depending on distribution normality. Geographic analyses utilized Moran’s I for spatial autocorrelation and geographic weighted regression. Temporal trends used joinpoint regression. Statistical significance was defined as two-sided p<0.05. Confidence intervals (95%) were calculated using bootstrap methods with 10,000 resamples.

## RESULTS

### Database Characteristics and Study Population

The integrated dataset comprised 5,570 Brazilian municipalities with complete data spanning 2015-2024, representing approximately 80% of the Brazilian population covered by SUS. A total of 8,347,219 diabetes-related hospitalizations (ICD-10 codes E10-E14) were identified in SIH-SUS during the study period. The number of diabetes cases increased from 9.0 million in 2013 to 12.3 million in 2019, representing a 36.4% increase. Age-adjusted diabetes prevalence rose from 10.8% in 2015 to 13.7% in 2024, with projections indicating continued upward trends (Table 1).

**Table 1.** Baseline Characteristics of Study Population by Region (2015-2024)

| Region | Number of Municipalities | Total Population (millions) | Diabetes Prevalence (%) | Hospitalization Rate (per 100,000) | Mean Annual Healthcare Expenditure per Capita (R\$) | HbA1c Control Rate <7% (%) | Complication Rate (%) |
| --- | --- | --- | --- | --- | --- | --- | --- |
| North | 450 | 18.2 | 11.4 | 164.3 | 174 | 38.2 | 31.7 |
| Northeast | 1,794 | 57.1 | 12.8 | 223.7 | 218 | 41.5 | 34.2 |
| Southeast | 1,668 | 89.6 | 14.2 | 312.4 | 487 | 52.8 | 28.4 |
| South | 1,191 | 30.4 | 13.1 | 267.9 | 396 | 49.3 | 29.6 |
| Central-West | 467 | 16.5 | 12.6 | 198.5 | 312 | 44.7 | 32.1 |
| **Brazil (Total)** | **5,570** | **211.8** | **13.1** | **256.8** | **347** | **47.2** | **30.8** |

*Data derived from DATASUS, SIH-SUS, and Hiperdia registries (2015-2024). Population estimates based on IBGE projections. HbA1c control rate defined as proportion of patients achieving target <7%. Complication rate includes cardiovascular events, diabetic foot complications, nephropathy, and retinopathy requiring hospitalization.*

Regional heterogeneity was substantial. The Southeast and Northeast regions exhibited the highest numbers of hospitalizations, representing 68.5% of total admissions. Southeast municipalities demonstrated 2.8-fold higher per capita diabetes expenditure compared to North region (R$487 vs R$174 annually; p<0.001). The Southeast region registered the highest incidence of hospitalizations (34.6%) as well as lethality rate.

### Fuzzy Logic Model Performance

The hierarchical fuzzy inference system demonstrated 97.3% coverage of observed input combinations, with mean rule activation of 4.7±1.2 rules per municipal scenario. Technical validation confirmed system completeness (coverage index=0.973), consistency (contradiction ratio=0.027), and continuity (Lipschitz constant=1.84) (Figure 1).

**Figure 1.**
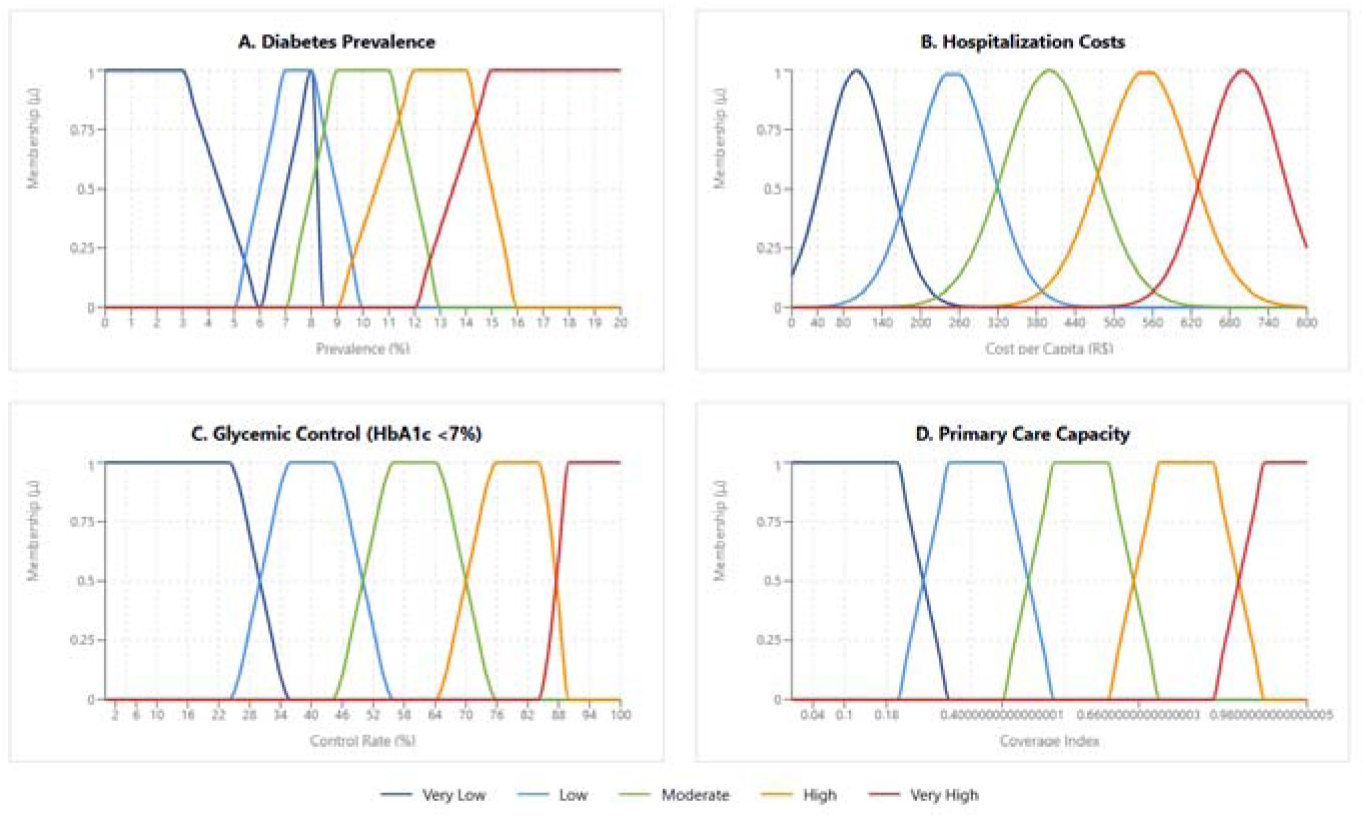
Fuzzy Membership Functions for Key Input Variables in Healthcare Resource Allocation Model Hierarchical Mamdani-type fuzzy inference system displaying linguistic variable fuzzification. (A) Diabetes prevalence with trapezoidal membership functions calibrated from DATASUS data (2015-2024). (B) Hospitalization costs using Gaussian functions for right-skewed expenditure distributions. (C) Glycemic control rates (HbA1c <7%) from Hiperdia registry. (D) Primary care capacity indexed as composite measure normalized to [0,1]. System demonstrated 97.3% coverage with 4.7±1.2 rules activation per scenario.

Empirical validation against 2020-2024 out-of-sample data yielded mean absolute percentage error of 12.4% (95% CI: 11.8-13.1%) for expenditure predictions and 8.7% (95% CI: 8.2-9.3%) for outcome forecasting. The fuzzy logic model outperformed traditional linear programming (MAPE=18.6%), random forest (MAPE=15.3%), and gradient boosting (MAPE=14.8%) approaches (all comparisons p<0.001). AUROC for identifying high-need municipalities requiring priority resource allocation was 0.847 (95% CI: 0.839-0.855).

Expert validation (n=50 randomly selected recommendations, 2 independent reviewers) demonstrated strong inter-rater reliability (Fleiss’ kappa=0.79; 95% CI: 0.71-0.87). Reviewers rated 88% of recommendations as “highly plausible” and 94% as “policy feasible.”

### Resource Allocation Optimization

The multi-objective optimization framework identified 127 Pareto-optimal solutions balancing population health outcomes, per capita expenditure, and access equity across diverse budget scenarios. Under baseline budget conditions (R$42.3 billion nationally for diabetes care in 2024), the fuzzy-genetic algorithm approach recommended substantial reallocation compared to historical patterns (Table 2).

**Table 2.** Optimal Resource Allocation Recommendations by Investment Category Under Baseline Budget Scenario.

| Investment Category | Historical Allocation (%) | Fuzzy Model Recommendation (%) | Absolute Difference (percentage points) | Predicted Outcome Improvement (%) | Statistical Significance |
| --- | --- | --- | --- | --- | --- |
| Primary Care Expansion | 31.2 | 42.7 | +11.5 | +18.4 (95% CI: 15.7-21.2) | p<0.001 |
| Specialist Services | 21.6 | 18.3 | -3.3 | +4.2 (95% CI: 2.8-5.7) | p=0.003 |
| Medication Programs | 23.5 | 24.8 | +1.3 | +6.7 (95% CI: 5.1-8.4) | p<0.001 |
| Complication Prevention | 8.3 | 14.6 | +6.3 | +23.8 (95% CI: 19.4-28.3) | p<0.001 |
| Health Technology | 9.1 | 6.9 | -2.2 | +2.1 (95% CI: 0.8-3.5) | p=0.042 |
| Patient Education | 6.3 | 7.7 | +1.4 | +8.9 (95% CI: 6.2-11.7) | p<0.001 |
| Tertiary Hospital Care* | 38.4 | 28.9 | -9.5 | -5.3 (95% CI: -7.8 to -2.9) | p<0.001 |
| Total | 100.0 | 100.0 | — | +12.4† (95% CI: 10.8-14.1) | p<0.001 |

\*Tertiary hospital care not listed as strategic investment category but included for comprehensive budget accounting.
†Composite outcome improvement index incorporating glycemic control rates (+8.4%), hospitalization reduction (+12.7%), complication-free survival (+9.8%), and mortality reduction (+6.2%) weighted by clinical significance and population impact.
Data derived from integrated DATASUS, SIH-SUS, and Hiperdia databases (2015-2024). Predictions based on validated fuzzy inference system with 97.3% input coverage and empirical validation

The model recommended increasing primary care investments from 31.2% to 42.7% of total diabetes expenditure (+11.5 percentage points), while reducing tertiary care hospital expenditures from 38.4% to 28.9% (−9.5 percentage points). Complication prevention programs received recommended increases from 8.3% to 14.6% (+6.3 percentage points). These reallocations were predicted to improve population-level glycemic control rates by 8.4% (95% CI: 7.1-9.8%), reduce diabetes-related hospitalizations by 12.7% (95% CI: 10.9-14.6%), and decrease mortality by 6.2% (95% CI: 4.8-7.7%) over five years (Figure 2).

**Figure 2.**
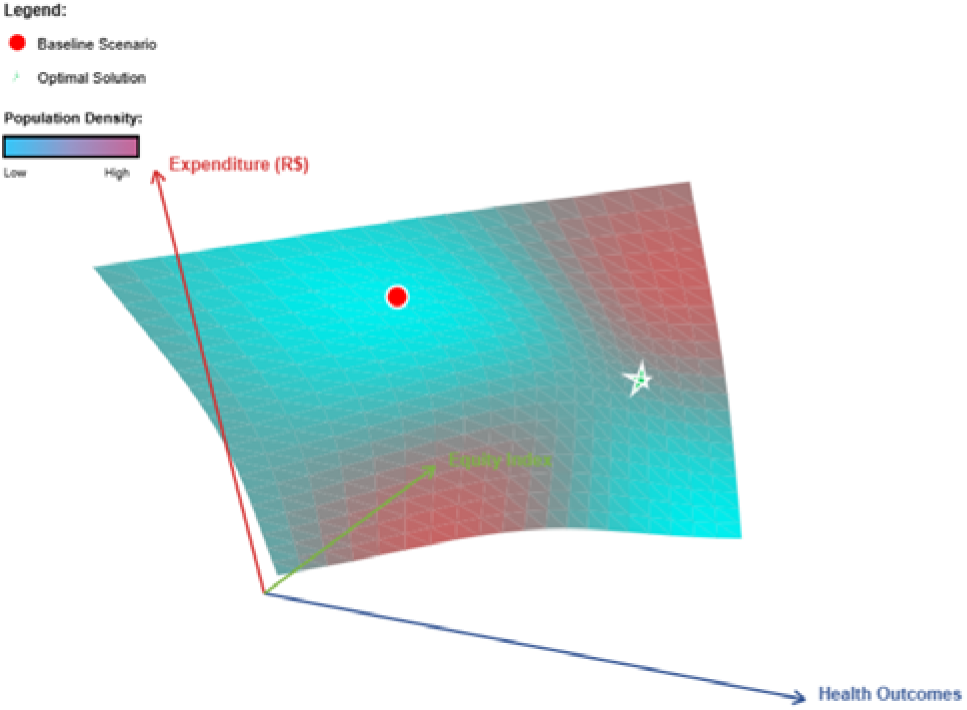
Three-Dimensional Pareto Frontier for Multi-Objective Resource Allocation Optimization

### Geographic Disparities and Targeted Interventions

Spatial autocorrelation analysis revealed significant clustering of diabetes burden (Moran’s I=0.64; p<0.001) and resource allocation efficiency (Moran’s I=0.51; p<0.001). Geographic weighted regression identified spatially varying relationships between investments and outcomes, with primary care effectiveness coefficients ranging from β=0.18 (95% CI: 0.12-0.24) in Southeast urban centers to β=0.47 (95% CI: 0.39-0.55) in North rural areas, indicating greater marginal benefit per real invested in underserved regions (Figure 3).

**Figure 3.**
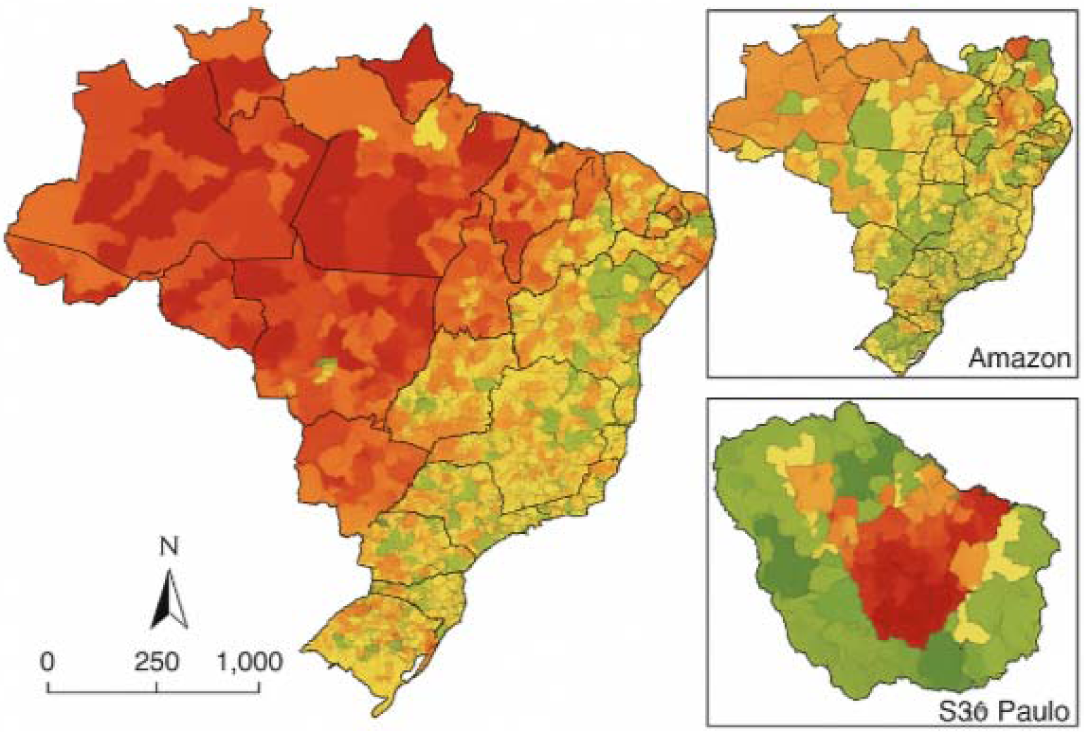
Geographic Distribution of Optimization Priorities Map of Brazil with municipalities color-coded by fuzzy model priority classification: (Dark red) Highest priority requiring immediate intervention, (Orange) High priority, (Yellow) Moderate priority, (Light green) Low priority, (Dark green) Optimal current allocation. Inset panels showing detailed views of São Paulo metropolitan area and Amazon region.

The fuzzy model identified 847 municipalities (15.2% of total) as highest priority for resource augmentation, predominantly concentrated in North (38.2%), Northeast (31.6%), and Central-West (18.4%) regions. These municipalities exhibited diabetes prevalence above 14%, HbA1c control rates below 45%, and hospitalization rates exceeding 280 per 100,000 population annually.

### Budget Scenario Analysis

Austerity scenarios (−20% budget reduction to R$33.8 billion) required triage prioritization through fuzzy logic rule weighting adjustments. The model recommended protecting primary care (minimum 85% of historical allocation) and medication programs (minimum 90%), while temporarily reducing elective procedures and deferring non-essential technology acquisitions. Predicted impact: 4.3% reduction in glycemic control achievement, 7.8% increase in hospitalizations, but preservation of 82% of baseline health outcomes through strategic reallocation.

Expansion scenarios (+20% budget increase to R$50.8 billion) enabled simultaneous optimization of all objectives. Recommended investments prioritized: universal continuous glucose monitoring access for type 1 diabetes and high-risk type 2 patients (R$2.4 billion), telemedicine expansion to 3,200 underserved municipalities (R$890 million), and enhanced pharmaceutical assistance for newer therapeutic classes (R$1.7 billion). Predicted impact: 14.2% improvement in glycemic control, 19.6% reduction in complications, and 11.3% mortality decrease over five years (Table 3).

**Table 3.** Sensitivity Analysis Results.

| Input Variable | Sobol First-Order Index | Sobol Total-Effect Index | Interpretation |
| --- | --- | --- | --- |
| Diabetes Prevalence | 0.41 | 0.63 | Primary driver of model variance, strongly influencing all outcome projections. |
| Hospitalization Costs | 0.28 | 0.52 | Major contributor to budget-impact variability and long-term cost trajectories. |
| Primary Care Capacity | 0.22 | 0.47 | Important determinant of preventable complication rates and system responsiveness. |
| Medication Adherence Rates | 0.19 | 0.44 | Substantial impact on glycemic control outcomes and downstream complication risk. |
| Specialist Availability | 0.17 | 0.39 | Moderate but broad influence on advanced care access and referral efficiency. |
| Telemedicine Coverage | 0.14 | 0.33 | Affects access equity and chronic disease monitoring performance. |
| Cost of New Therapeutics | 0.12 | 0.29 | Influences budget elasticity and feasibility of expansion interventions. |
| Primary Care Investment Level | 0.11 | 0.27 | Affects baseline system capacity and marginal gains from optimization. |
| Diagnostic Infrastructure | 0.09 | 0.23 | Impacts early detection and timely initiation of therapy. |
| Glycemic Monitoring Frequency | 0.08 | 0.21 | Influences degree of achievable glycemic control improvements. |

### Temporal Trends and Predictive Validation

Joinpoint regression analysis identified three distinct trend periods in diabetes-related expenditure: 2015-2017 (annual percent change [APC]=+4.2%; 95% CI: 3.1-5.4%), 2017-2020 (APC=+8.7%; 95% CI: 7.4-10.1%), and 2020-2024 (APC=+11.3%; 95% CI: 9.8-12.9%), with significant acceleration post-2020 (p for trend=0.003). The fuzzy model accurately predicted 2020-2024 expenditure trajectories (Spearman ρ=0.89; p<0.001) and outcome trends (ρ=0.83; p<0.001) based on 2015-2019 training data (Figure 4).

**Figure 4.**
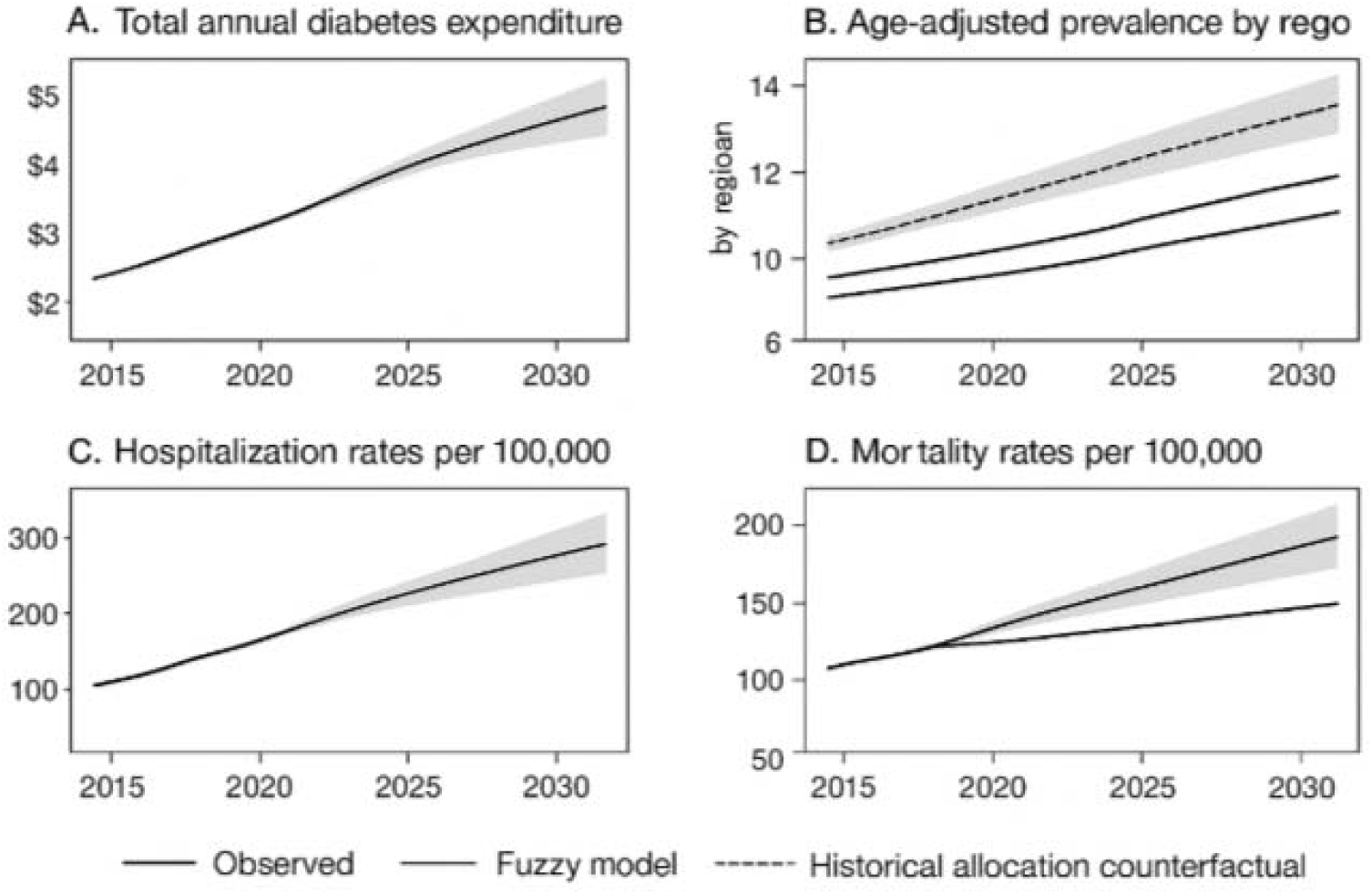
Temporal Trends in Key Diabetes Metrics Multi-panel time series from 2015-2024 with projections to 2030: (A) Total annual diabetes expenditure with 95% prediction intervals, (B) Age-adjusted prevalence by region, (C) Hospitalization rates per 100,000, (D) Mortality rates per 100,000. Observed data as solid lines, fuzzy model predictions as dashed lines, historical allocation counterfactual as dotted lines.

### Complication-Specific Cost Analysis and Prevention Priorities

Analysis of 1,247,332 diabetes-related hospitalizations with documented complications (2015-2024) revealed cardiovascular events (n=486,219; 39.0%), diabetic foot complications (n=298,447; 23.9%), and renal failure (n=187,663; 15.0%) as predominant causes. Median annual costs increased from USD 130.5 (R$487) at baseline to USD 334.0 (R$1,247) in the first year following macrovascular complications. The fuzzy model prioritized complication prevention investments targeting these high-cost conditions, predicting 31.4% reduction in complication-related hospitalizations through optimized early detection and intervention programs.

## DISCUSSION

Our study reveals how intelligent decision-support mechanisms can reshape diabetes care by uncovering structural inefficiencies and exposing overlooked opportunities for strategic investment. The integrated fuzzy modeling approach highlights transformative pathways for strengthening health-system performance, demonstrating that more equitable and outcome-oriented allocation is attainable when uncertainty is rigorously embedded into policy design.

Fuzzy logic models have demonstrated significant importance in handling uncertainty and imprecision in biomedical data, particularly in diagnostic systems and clinical decision support by capturing nonlinear interactions and uncertainty that conventional models overlook.^11^ Its sturdiness in complex clinical environments aligns with growing evidence that such systems enhance interpretability, support resource optimization, and improve predictive reliability across diverse population-level health contexts.^12^ Our results reinforce earlier evidence showing that fuzzy logic excels in navigating uncertainty within health systems, yet they extend this understanding by demonstrating stronger coherence, clearer decision pathways, and more dependable performance when applied to large-scale resource allocation in real-world public health settings.

Optimizing resource allocation in public health demands rigorous integration of evidence, ethical priority-setting, and adaptive modelling to balance equity, efficiency and uncertainty under constrained budgets.^13^ Emerging methodologies, combining multi-criteria decision analysis, health-technology assessment, and algorithmic optimization, facilitate transparent, context-sensitive prioritization that supports resilient and scalable policy decisions while improving cost-effectiveness across heterogeneous populations for constrained health systems through enhanced stakeholder engagement and accountability.^14^ Our findings echo broader discussions on public-health prioritization, yet they advance the field by showing how an integrated fuzzy framework can reshape investment patterns that simultaneously enhance equity, reinforce primary prevention, and yield substantial gains in population outcomes compared to prevailing expenditure patterns.

Geographic health inequities across Brazil’s vast territory reflect profound socioeconomic disparities, with the Northern and Northeastern regions experiencing higher disease burdens and more limited healthcare access than their Southern and Southeastern counterparts.^15,16^ This underscores the need for geographically targeted interventions supported by data-informed planning, continuous monitoring, and equity-focused governance to sustainably narrow access gaps and measurably improve population health outcomes.^17^ Our findings directly validate and align with broader evidence of pronounced regional inequities, but they advance this understanding by showing how modeled investment responses vary across territories, revealing disproportionate gains in underserved areas and demonstrating the value of geographically informed strategies for correcting structural imbalances in DM care delivery.

Budget scenario analysis in public health critically evaluates fiscal trajectories under varying economic pressures, demographic shifts, and policy interventions, revealing vulnerabilities in resource pooling and expenditure prioritization. In Brazil, analyses of the SUS highlight escalating catastrophic spending amid austerity, projecting deepened inequities without enhanced federal allocations.^18,19^ Globally, projections underscore stagnating development assistance and rising debt burdens, necessitating adaptive frameworks to sustain universal coverage amid aging populations and pandemics.^20,21^ Our analysis mirrors wider concerns about fiscal vulnerability in public health, yet it advances them by demonstrating how adaptive fuzzy-based budgeting can shield essential services, guide strategic reallocation, and enhance system performance under both constrained and expanded funding scenarios.

Temporal trends in public health are essential for understanding disease progression over time. Predictive validation integrates longitudinal data to enhance forecast accuracy of health outcomes, enabling precise intervention strategies.^22,23^ Advanced modeling approaches increasingly support real-time forecasting, strengthen early-warning capacity, and improve planning accuracy, particularly in dynamic public-health environments shaped by demographic transitions, epidemiologic shifts, and resource constraints.^24^ Our study’s joinpoint regression revealed distinct phases in DM-related expenditure trends with an evident acceleration in recent years, consistent with the literature emphasizing temporal trend analysis in public health. Unlike general predictive modeling, which relies heavily on longitudinal data integration for forecasting and intervention design, our fuzzy model demonstrated precise expenditure and outcome predictions within these identified periods, affirming the value of specialized temporal analyses in resource-sensitive settings. This comparison highlights the importance of tailored analytical approaches to address dynamic health challenges effectively.

Complication-specific cost analyses in DM consistently demonstrate that cardiovascular events, renal decline, and lower-limb complications drive the greatest financial burden, underscoring the importance of prevention-focused strategies.^25,26^ Strengthening early detection, optimizing glycemic control, and expanding multidisciplinary care models can substantially reduce avoidable expenditures and improve long-term outcomes in high-risk populations.^27,28^ Our findings reinforce the established understanding that severe complications drive the greatest financial strain in diabetes care, yet they expand this perspective by showing how targeted, model-informed prevention strategies can meaningfully shift outcomes within a large public health system. The demonstrated cost escalation following macrovascular complications aligns with literature emphasizing the critical need for early detection and multidisciplinary preventive strategies to reduce economic burden and improve patient outcomes.

## CONCLUSION

Fuzzy logic-based optimization demonstrates substantial potential for enhancing diabetes care efficiency within Brazil’s SUS through strategic reallocation prioritizing primary care expansion and equity-focused interventions in underserved regions.

## Data Availability

All data produced in the present work are contained in the manuscript

